# Genome sequencing of sewage detects regionally prevalent SARS-CoV-2 variants

**DOI:** 10.1101/2020.09.13.20193805

**Authors:** Alexander Crits-Christoph, Rose S. Kantor, Matthew R. Olm, Oscar N. Whitney, Basem Al-Shayeb, Yue C. Lou, Avi Flamholz, Lauren C. Kennedy, Hannah Greenwald, Adrian Hinkle, Jonathan Hetzel, Sara Spitzer, Jeffery Koble, Asako Tan, Fred Hyde, Gary Schroth, Scott Kuersten, Jillian F. Banfield, Kara L. Nelson

**Affiliations:** Department of Plant and Microbial Biology, University of California, Berkeley, CA, USA; Innovative Genomics Institute, Berkeley, CA, 94704, USA.; Department of Civil and Environmental Engineering, University of California, Berkeley, CA, USA; Department of Microbiology and Immunology, Stanford University, CA, USA; Department of Molecular and Cell Biology, University of California, Berkeley, CA, USA; Illumina, San Diego, CA, USA; Department of Environmental Science, Policy, and Management, University of California, Berkeley, CA, USA; Earth Sciences Division, Lawrence Berkeley National Laboratory, Berkeley, CA, USA; Chan Zuckerberg Biohub, San Francisco, CA, USA

## Abstract

Viral genome sequencing has guided our understanding of the spread and extent of genetic diversity of SARS-CoV-2 during the COVID-19 pandemic. SARS-CoV-2 viral genomes are usually sequenced from nasopharyngeal swabs of individual patients to track viral spread. Recently, RT-qPCR of municipal wastewater has been used to quantify the abundance of SARS-CoV-2 in several regions globally. However, metatranscriptomic sequencing of wastewater can be used to profile the viral genetic diversity across infected communities. Here, we sequenced RNA directly from sewage collected by municipal utility districts in the San Francisco Bay Area to generate complete and near-complete SARS-CoV-2 genomes. The major consensus SARS-CoV-2 genotypes detected in the sewage were identical to clinical genomes from the region. Using a pipeline for single nucleotide variant (SNV) calling in a metagenomic context, we characterized minor SARS-CoV-2 alleles in the wastewater and detected viral genotypes which were also found within clinical genomes throughout California. Observed wastewater variants were more similar to local California patient-derived genotypes than they were to those from other regions within the US or globally. Additional variants detected in wastewater have only been identified in genomes from patients sampled outside of CA, indicating that wastewater sequencing can provide evidence for recent introductions of viral lineages before they are detected by local clinical sequencing. These results demonstrate that epidemiological surveillance through wastewater sequencing can aid in tracking exact viral strains in an epidemic context.

## Introduction

The COVID-19 pandemic caused by SARS-CoV-2 reached the United States at the start of 2020, with multiple early introduction events in the states of Washington, California, and New York [1]. Since then, the total number of cases in the country has surpassed 6 million, with over 180,000 deaths and enormous implications for public health [2]. While clinical viral cases have been tracked mostly with quantitative reverse transcriptase PCR (RT-qPCR), there has also been extensive whole viral genome sequencing of clinical cases, generating over 75,000 genomes globally, including 17,000 from the US, and 2,500 from California (GISAID EpiCov database as of August 23, 2020)[3].

Genomic epidemiology, the analysis of viral genomes in order to make inferences about viral evolution, transmission, and spread, has played an important role in improving our understanding of the transmission dynamics of the SARS-CoV-2 pandemic [4]. Early in the pandemic, this approach revealed multiple introduction events into California and viral lineages present at different abundances across counties in Northern California [5]. Genome sequencing was also used to show that there was unexpectedly frequent community spread of a specific genotype after early introduction in Washington State [6]. Genome sequencing in the New York City area identified multiple viral introduction events from Europe [7] and sequencing in the Mission district of San Francisco identified distinct viral strains in a single neighborhood, with transmission between family clusters [8].

Unlike many respiratory viruses, RNA of SARS-CoV-2 and other coronaviruses can be detected in human feces [9–11]. Before the COVID-19 pandemic, members of the coronaviridae had been previously identified in municipal wastewater through both RT-qPCR and shotgun metagenomic and metatranscriptomic sequencing [12,13]. Since the start of the COVID-19 pandemic, wastewater RT-qPCR has quantified the amount of SARS-CoV-2 RNA in sewage to estimate the abundance of the virus across many different municipal regions globally [14–22]. Prior work showed that shotgun wastewater sequencing can provide information about many viruses simultaneously [12,23,24] and enable genome-resolved [25] and phylogenetic analyses [26,27]. In one study, a SARS-CoV-2 consensus genome was obtained from sewage via targeted amplification and long-read sequencing, allowing for phylogenetic analysis of the predominant lineage [27]. Here, we show that sequencing of viral concentrates and RNA extracted directly from wastewater can identify multiple SARS-CoV-2 genotypes at varying abundances known to be present in communities, as well as additional genotypic variants not yet observed in local clinical sequencing efforts.

## Results and Discussion

### Metatranscriptomic detection of SARS-CoV-2 and other viruses in wastewater

Twenty-four-hour 1L composite samples of raw sewage were collected from wastewater treatment facilities in Alameda and Marin Counties in Northern California between May 19, 2020 and July 15, 2020 (**Supplementary Table S1**). We extracted nucleic acids from samples using three methods that enriched for viral particles (ultrafiltration) or total RNA (RNA silica columns or silica milk). SARS-CoV-2 viral RNA was first detected using a RT-qPCR assay (see *Methods*) of the N gene and Cq-values ranged from 29.5 to 36.2, or an estimated ∼2 to ∼553 genome copies/μL of RNA. From this we estimate that there were 2.8×10^5^ genome copies/L of wastewater on average across our samples (**Supplementary Table S1**). For each sample, 40–50 μL of RNA was prepared for sequencing, implying an estimated ∼4438 viral genome copies on average were contained within each sequencing library.

After cDNA synthesis from the total RNA, samples were enriched for a panel of human respiratory viruses using a commercially available oligo-capture approach (Illumina Respiratory Virus Panel; see *Methods*)and sequenced on a NextSeq 550 to produce on average 12 million 2×75 bp reads per sample. Reads were mapped to the human genome to estimate the amount of human RNA/DNA in the samples (0.7–16% of reads per sample). Sequencing reads were then mapped to a de-replicated set of all eukaryotic viruses contained in the RefSeq database, and stringently filtered to include only high-quality reads matching reference sequences with >97% identity (*Methods*). Viral abundances and SNVs (Single Nucleotide Variants) were then calculated using the metagenomic strain-typing program inStrain v1.12. We detected SARS-CoV-2 at varying abundances (0%-14%) across samples (**
Fig 1a and 1b**; **Supplementary Table S1**). Sequencing relative abundance of SARS-CoV-2 was not strongly correlated with RT-qPCR genome copy quantification, likely due to the variability introduced by different extraction methods. Viral enrichment by ultrafiltration achieved higher relative abundances of SARS-CoV-2 RNA, although these experiments were time-intensive and often had lower absolute genome copy number recovery according to RT-qPCR. Additionally, we sequenced replicates from one set of samples with rRNA depletion but no viral enrichment. Without enrichment, we were able to only detect fewer than 40 total SARS-CoV-2 read pairs (**Supplementary Table S1**; **Fig 1c**). While this illustrates the difficulty of detecting specific viruses in wastewater in unenriched sequencing datasets, larger sequencing efforts may overcome this limitation by sequencing more deeply.

**Figure 1:**
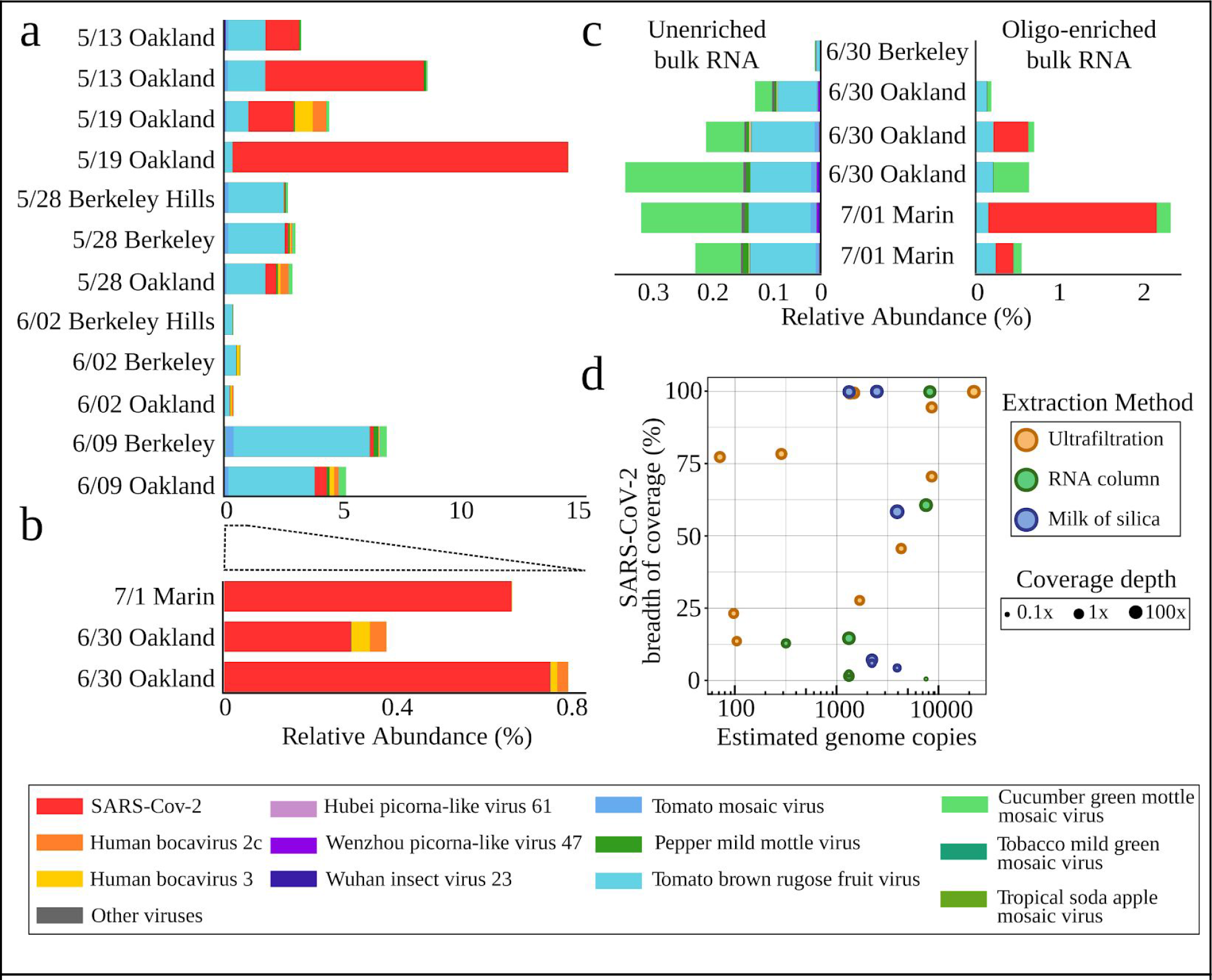
Characterized viruses detected in enriched and unenriched wastewater metatranscriptomes. Relative abundances of viruses with eukaryotic hosts in the RefSeq database as a percentage of total sequencing reads derived from the sample in **(a)** amicon ultrafiltration (viral fractionation) and **(b)** total RNA column and milk of silica samples. All samples were enriched with the Illumina Respiratory Virus Panel. **(c)** Relative abundances of RefSeq viruses in unenriched metatranscriptomics (left) and the same samples after oligo-enrichment with the Illumina Respiratory Virus Panel. **(d)** The relationship between the quantity of viral genome copies in 40 μL of purified RNA and SARS-CoV-2 genome completeness (measured in breadth of coverage) for each sample. Samples are colored by extraction methodology, and the size of the point corresponds to the mean SARS-CoV-2 depth of coverage.

Other human viruses identified in the wastewater sequencing included Human bocaviruses 2c and 3 (**
Fig 1a, 1b**), both of which are respiratory viruses sometimes capable of causing gastroenteritis, and are included in the Illumina Respiratory Virus Panel. Bocaviruses have been identified in sewage samples previously [28,29]. Picorna-like viruses were also detected (**Fig 1**). The most abundant viruses in the data were plant viruses including cucumber green mottle mosaic virus and pepper mild mottle virus (PMMoV) (**
Fig 1a
**). These viruses are known to be highly abundant in human wastewater [30] and have been used as fecal loading controls in wastewater SARS-CoV-2 quantification [19]. Near-complete (>95% breadth of coverage) genomes were obtained for SARS-CoV-2, bocavirus 3, PMMoV, and other plant viruses (**Supplementary Table S2**), implying that these viruses were at high enough abundance in the dataset for exact genomic analysis.

### Recovery of complete and near-complete SARS-CoV-2 viral genomes from wastewater

Complete consensus viral genomes are required to perform viral lineage tracking for genomic epidemiology. We obtained complete consensus SARS-CoV-2 genomes (breadth of coverage > 99%) from 7 out of 22 samples (31%), while large-scale patient sequencing efforts have for example obtained genomes for ∼80% of samples [31]. Only samples with RT-qPCR Ct-values < 33 (∼25 gc/uL) yielded complete consensus genomes (**
Fig 1d
**), but we also recovered at least one genome using each of our three extraction methods. Mean depth of coverage for each complete genome ranged from 7x to 107x after filtering and removal of PCR duplicates. The consensus genomes from Alameda County, and the one from Marin County, were all within 4 base pair differences of each other. These consensus genomes were found to be unlikely to be chimeric, as a BLAST analysis identified SARS-CoV-2 genomes that were 100% identical at all non-gapped positions (**Supplementary Table S3**) obtained from patients in Northern California. Consensus genomes may represent predominant SARS-CoV-2 lineages in the population in the serviced areas during the summer of 2020. The results demonstrate genomic accuracy for recovery of consensus SARS-CoV-2 genomes so long as sufficient coverage is achieved in metatranscriptomic datasets.

### Identification of alternative SARS-CoV-2 variants in wastewater populations recovers locally reported clinical genotypes

While consensus genotypes can describe the predominant genotype of a virus in a metatranscriptome, the strength of wastewater-based sampling and sequencing lies in the ability to identify alternative genotypes in the population being sampled. Using a recently developed pipeline for metagenomic SNV calling [32], we identified putative SNVs that are variable within the viral population sampled in each wastewater sample (**
Fig 2a; Supplementary Table S4**). Due to the large scale sequencing efforts of SARS-CoV-2 in patients in both Northern California and worldwide, we established that these SNVs had also been detected in genomes from individual patients. Across all samples, 50% of SNVs observed in wastewater samples at greater than 10% frequency were also observed in patient-derived viral genomes from California; 61% were observed in viral genomes from the USA, and 71% were observed in any viral genomes collected worldwide. SNVs that have been observed in California patients had significantly higher allele frequencies in the wastewater samples than those that were not detected in clinical cases (mean 48% versus 15%, respectively; p< 0.01; two-sided t-test) (**
Fig 2b
**). This is likely because the more abundant a SNV is in the population, the more likely it is to be sampled in wastewater and in the clinic. Further, several of the same SNVs were observed across samples, and these recurrent SNVs were on average 2.3x more likely to be observed in California or USA patient-derived genomes than SNVs observed once (**
Fig 2c
**). Taken together, these are strong signals that deeper sequencing of wastewater and combining information across samples better recapitulates true viral genomic variation in the sampled population.

**Figure 2:**
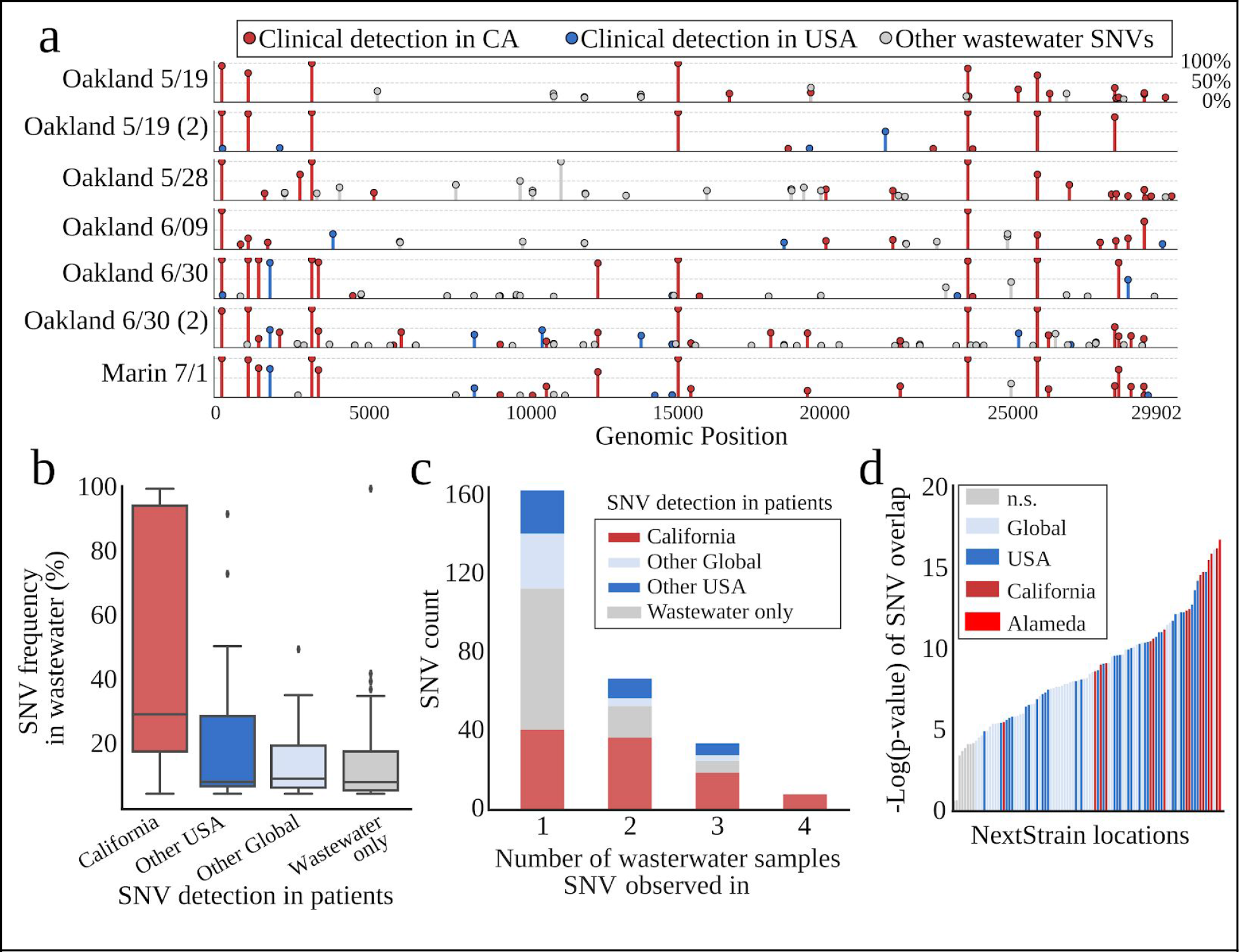
SARS-CoV-2 SNVs in wastewater samples. **(a)** Allele frequencies of SARS-CoV-2 in wastewater metatranscriptomes for each sample. Each point is a SNV by location on the SARS-CoV-2 genome (x-axis), and the height of the bar (y-axis) is the frequency of the alternative allele (relative to the reference genome EPI_ISL_402124) at that position. Wastewater SNVs are colored by whether they have previously been observed in clinical samples from CA, the USA, or neither. **(b)** Wastewater SARS-CoV-2 frequencies grouped by whether they have been observed in clinical samples from different regions. Most highly abundant SNVs have been observed previously in California or elsewhere in the US. **(c)** SARS-CoV-2 SNVs grouped by the number of wastewater samples observed in (out of 7 high quality samples). Most SNVs that were observed in 2 or more samples have been observed clinically in CA. **(d)** Multiple hypothesis adjusted (bonferroni correction) *p*–value distribution of hypergeometric tests for overlap between all wastewater SNVs observed and the variants clinically observed and reported in each location (a county level designation in the United States). Alameda County was the most significant comparison.

Over 75,000 patient-derived SARS-CoV-2 genomes have been sequenced and deposited into the GISAID database globally, including 2,500 genomes obtained from patients in California. To understand the context of the viral genomic variation we observed within wastewater samples, we used a hypergeometric test to calculate the likelihood of overlap by chance between the set of wastewater variants and the set of variants observed in viruses from patients in a given region. This computes the probability of observing a certain amount of overlap in variants by chance, and accounts for the fact that some regions have far more sequenced patient genomes and correspondingly more alleles than others. For example, the probability of the observed overlap between wastewater variants and California clinical variants having occurred by chance was calculated to be P< 10^−10^, indicating a high likelihood of non-random overlap. By further comparing the probabilities of SNV overlap between patient genotypes and wastewater genotypes at the nextStrain “location” level (corresponding to counties and/or cities), we found the highest likelihood of non-random overlap between all wastewater genotypes observed and clinical genotypes from Alameda County (**
Fig 2d
**) – the location that the wastewater samples were also derived from.

### Identification of potential lineage transmission events previously undetected in local patient-based sequencing at time of sampling

Some clinical SARS-CoV-2 viral strains can be differentiated by more than one SNV. Across the wastewater dataset, we observed one pair and one triplet of SNVs that were shared by clinical isolates. The pair and triplet of SNVs each occurred at similar frequencies, supporting their linkage in wastewater genomes (**
Fig 3a and 3b**). In addition to the SNVs that also have been observed clinically in California, there were four SNVs recurrent across wastewater samples that had not been previously observed in CA, but had been observed elsewhere in the United States (**Fig 3c**). Two adjacent SNVs (14222G and 14223C) are associated with a single viral strain that has been often observed in clinical samples in Washington State. Another two SNVs (8083A and 1738T) are not linked, but both have been observed in different clinical genomes of four other states in the US. Interestingly, these variants appear to have arisen or arrived in the US only during the month of July, suggesting that they may be detected in clinical samples from California in the near future.

**Figure 3:**
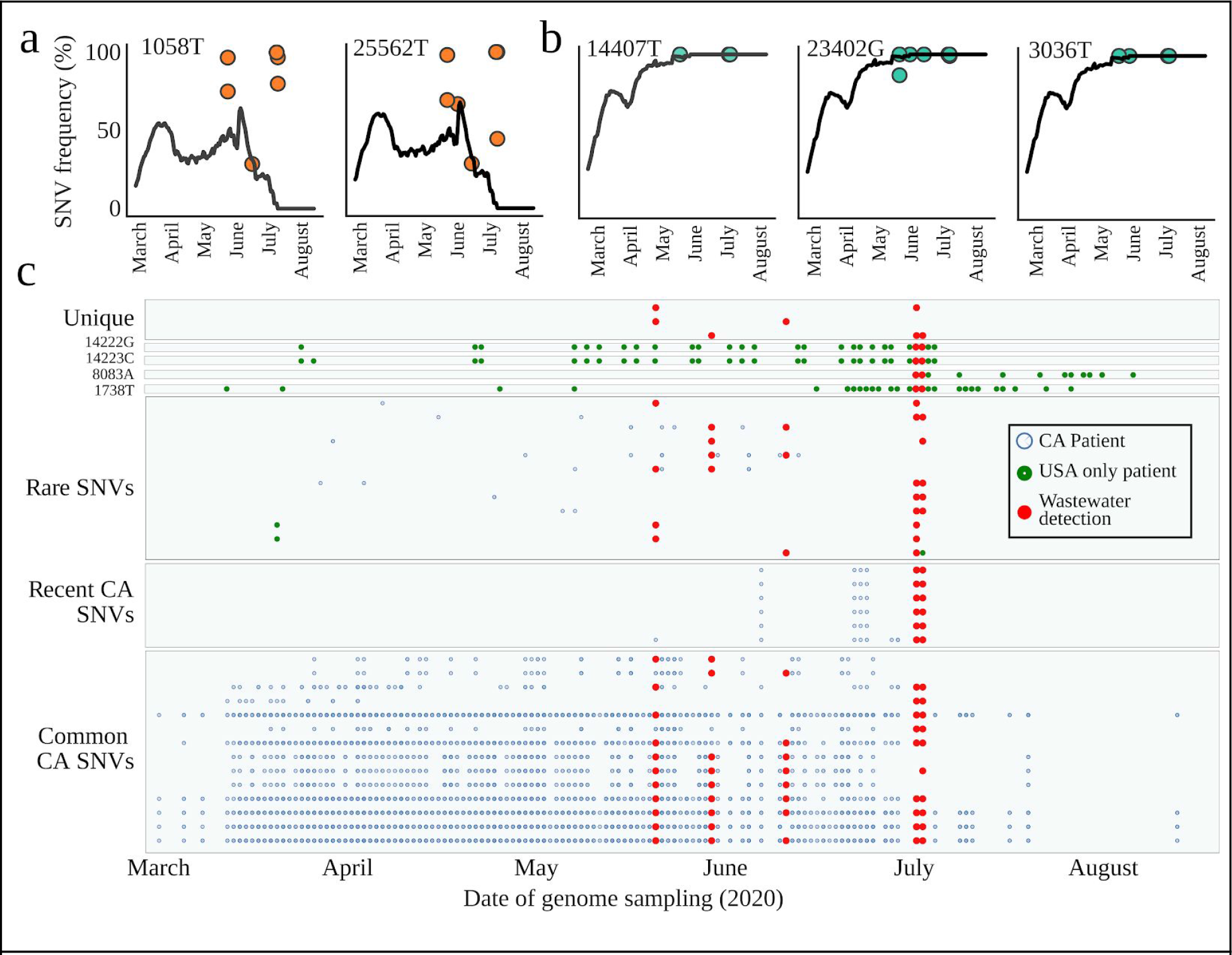
Time series of SARS-CoV-2 genotypes in California wastewater compared to patients. **(a)** Left two plots: Frequencies of two SNVs found in the same viral lineage across California clinical samples (black lines) and within each wastewater sample (orange points). **(b)** Frequencies of three SNVs found in the same viral lineage across California clinical samples (black lines) and within each wastewater sample (green points). **(c)** Time series of detection for recurrent wastewater genotypes in clinical samples versus wastewater samples. Each row on the y-axis is a SNV, and the presence of a point along the x-axis indicates when that SNV was detected in either a clinical sample or a wastewater sample.

Overall, this study demonstrated that wastewater sequencing can accurately identify genotypes of viral strains that are clinically detected in a region, and those not yet detected by clinical sequencing. Another key advantage of this method is that it does not rely on specific PCR primers, which can fail to detect SARS-CoV-2 strains with mutations in the primed sequence [33]. With more intensive wastewater sampling, this approach also has the potential to reveal patterns of virus distribution within communities, helping understand transmission and spread of diseases during epidemics. Perhaps most significantly, the results indicate that wastewater sequencing can detect recent introductions of SARS-CoV-2 genotypes and other disease-causing viruses at a population scale.

## Methods

### Sample collection and extraction

Twenty-four-hour 1L composite samples were collected at 4 different wastewater interceptors in the San Francisco Bay Area (labeled ‘Berkeley’, ‘Berkeley Hills’, ‘Oakland’, and ‘Marin’, based roughly on the municipal areas each services). Samples were immediately processed by extraction via three different methods. The first method was ultrafiltration with Amicon Ultra-15 100 kDa Centrifugal Filter Units. Wastewater was heat inactivated in a water bath at 60 degrees C for 90 minutes. Wastewater samples were then filtered on 0.22 µM SteriFlip filter units. Amicon filter units were prepared by incubation with 1% Bovine Serum Albumin in 1x phosphate buffered saline (PBS) on ice for 1 hour, after which they were spun, loaded with 2 mL PBS, and spun again to rinse. Amicon 100 kDa centrifugal filter units were then loaded with 15 mL of filtered wastewater and spun in a swinging bucket rotor at 4750 g for 30 min at 4C. Flow-through was discarded and amicons were reloaded with sample until all sample volume (40 mL) had been processed. For three samples (**Supplementary Table S1**), we processed more than 40 mL per sample, but found that this did not improve resulting SARS-CoV-2 genome quality in this specific instance. For all amicon-concentrated samples, the final volume of the concentrate was ∼250 μL. RNA was then extracted with a Qiagen AllPrep DNA/RNA Mini Kit. The second extraction method, direct RNA extraction with silica columns, began with viral and bacterial lysis of samples with 9.5 g of NaCl per 40 mL of wastewater and filtration on a 5 µM Polyvinylidene Fluoride (PVDF) filter. Resulting lysate was then loaded onto a Zymo III-P silica spin column via vacuum manifold, and RNA was directly eluted from this column. Details of this protocol are available at https://www.protocols.io/view/v-2-direct-wastewater-rna-capture-and-purification-bjr9km96. The third extraction method, “milk of silica”, began with sample lysis and filtration, as in the second method. Filtered lysate is bound to free silicon dioxide particulate, eluted from the particulate, and concentrated via isopropanol precipitation. This protocol is available at: https://www.protocols.io/view/direct-wastewater-rna-extraction-via-the-34-milk-o-biwfkfbn.

### RT-qPCR and genome copy quantification

The number of viral genome copies in each sample was determined via qRT-PCR on an Applied Biosystems QuantStudio 3 Real-Time PCR System with the Thermo Fisher TaqPath 1-Step RT-qPCR Master Mix or TaqMan™ Fast Virus 1-Step Master Mix. The primer set was purchased as part of the 2019-CoV RUO Kit (IDT) and our quantification used the previously published CDC N1 assay [34]. Either 2 μL or 5 μL of sample were used for each reaction (**Supplementary Table S1**) in a 10 μL or 20 μL reaction, respectively. Cycling conditions were 25 °C for 2 minutes, 50 °C for 15 minutes, 95 °C for 2 minutes, and 45 cycles of 95 °C for 3 seconds, 55 °C for 30 seconds. A standard curve for absolute quantification of viral genome copies was generated with synthetic RNA standards of the SARS-CoV-2 genome (Twist Biosciences).

### Library preparation and sequencing

Sequencing for a first set of samples was performed at the Microbial Genome Sequencing Center (Pittsburgh, PA) in three independent sequencing runs. The Maxima ds cDNA RT kit (Thermo Fisher) was used to generate cDNA. The Illumina Flex for Enrichment kit paired with the Illumina Respiratory Virus Oligo Panel (Illumina, Inc.) were used to enrich for respiratory virus cDNA with 15 PCR cycles in the final step. The libraries were then sequenced on a NextSeq 550 to yield on average 119 Mbp of 2×75 bp paired end sequencing reads. For a second set of samples (**Supplementary Table S1**), rRNA depletion was performed and oligo-capture enriched and unenriched sequencing strategies were compared. The rRNA depletion was done using RiboZero Plus supplemented with a comprehensive ‘Gut Microbiome’ probe set. Libraries were prepared using the Illumina RNA Prep with Enrichment (L) Tagmentation protocol. The rRNA depleted samples were amplified for 20 cycles. Enrichment was performed using the Illumina Respiratory Virus Oligo Panel.

### Metatranscriptomic viral abundances

The abundances of viruses within wastewater were obtained by mapping reads with Bowtie 2 [35] to an index of all viral genomes downloaded from the RefSeq Database (Release 201). For abundance calculations, mapped read pairs with MAPQ>20 and pair percent identity to the reference >95% were retained using inStrain v1.3.2 [32]. Duplicate reads were removed with the clumpify.sh dedup command from the BBTools software suite (Bushnell 2014). Only viral genomes with at least 10% breadth of genomic coverage obtained were reported.

### SARS-CoV-2 variant analysis

Seven samples with near-complete SARS-CoV-2 breadth of genomic coverage (>99%) were further investigated for a strain-resolved analysis. SNV calling was performed using inStrain v1.3.2 on all read pairs with >90% Average Nucleotide Identity to the SARS-CoV-2 reference. An absolute minimum of 2 read pairs supporting a variant allele was required for any SNV to be considered in further analysis. PCR duplicates were removed with the markdup command in the Sambamba package [36]. All analysis and SNV locations reported are with respect to the reference genome “hCoV-19/Wuhan/WIV04/2019|EPI_ISL_402124 |2019–12–30|China”. Consensus genomes from each sample were created using a custom Python script that required a minimum of 3 reads supporting each genomic position. A multiple sequence alignment of publicly available SARS-CoV-2 genomes and their metadata were downloaded from the GISAID [3] EpiCov database on August 23, 2020. The multiple sequence alignment was processed with a custom Python script to obtain a list of variants for each genome with respect to the WIV04 reference sequence. We removed from all analyses the genomic positions recommended to be masked from SARS-CoV-2 alignments by https://virological.org/t/masking-strategies-for-sars-cov-2-alignments/480. Hypergeometric distributions were calculated with the stats.hypergeom function in scipy [37] to compare wastewater samples to all clinical data from each NextStrain “location” with at least 20 genomes deposited. The following parameters were used for hypergeometric distribution testing: the total number of SNVs observed across all clinical SARS-CoV-2 genomes, the number of SNVs observed in wastewater, the number of clinical SNVs in a region, and the observed overlap between the two. Reproducible code is available at https://github.com/alexcritschristoph/wastewater_sarscov2.

## Data Availability

Sequencing data for this project has been released under NCBI BioProject ID PRJNA661613. Processed data, reproducible code, and workflows for the analyses performed have been made available at https://github.com/alexcritschristoph/wastewater_sarscov2.

https://www.ncbi.nlm.nih.gov/bioproject/PRJNA661613

## Acknowledgements

We gratefully acknowledge the originating and submitting laboratories of SARS-CoV-2 genomes in the GISAID EpiCoV database (https://www.gisaid.org) that were used for our comparisons to clinical samples and in particular the Innovative Genomics Institute SARS-CoV-2 Sequencing Group for Alameda County genomes. We also gratefully acknowledge Vinson Fan for assistance with RT-qPCR and the laboratory of Robert Tjian for sharing materials. Funding was provided to KLN and JFB by a Rapid Research Response grant from the Innovative Genomics Institute (IGI) and a seed grant from the Center for Information Technology Research in the Interest of Society (CITRIS) at UC Berkeley.

